# Body Image Group as Adjunct to Eating Disorder Treatment: Feasibility, Acceptability, and Early Outcomes

**DOI:** 10.1101/2025.05.06.25327081

**Authors:** Catherine R. Drury, Erin E. Reilly, Naomi Lynch, Erin C. Accurso, Kathryn M. Huryk

**Author notes:** Correspondence concerning this article should be addressed to Catherine R. Drury, Department of Psychiatry and Behavioral Sciences, University of California, San Francisco, 675 18^th^ Street, San Francisco, CA 94143, United States.

## Abstract

Negative body image (BI) contributes to the development and persistence of eating disorders (EDs) and is linked to poorer treatment outcomes and higher risk of relapse. However, existing ED treatments show limited effectiveness in reducing negative BI, often compelling psychotherapists to integrate additional BI techniques into treatment to address their patients’ concerns. This study evaluates the feasibility and preliminary outcomes of a 12-session virtual BI group intervention, designed to programmatically integrate evidence-based BI protocols into ED treatment in response to this clinical need. Thirty-six adolescents and young adults (age range: 14 to 21 years) with transdiagnostic EDs enrolled in the group through an outpatient ED clinic, where they were already receiving specialized individual and/or family therapy. Participants were required to be willing to participate in group BI discussions, and their weight needed to be >90% of their expected body weight. Before and after group participation, participants were asked to complete an online survey that included measures of group acceptability, negative body image, and overall eating disorder pathology. Preliminary findings are promising for the feasibility, acceptability, and effectiveness of this intervention. Most referrals enrolled and successfully engaged, with 63.9% having completed or still actively participating in the group at the end of the data collection period and demonstrating reductions in negative BI and overall ED pathology. Participants’ qualitative feedback emphasized the importance of adapting BI interventions for all genders and body sizes. Additional research is needed to experimentally test the BI group’s effectiveness in larger samples and other treatment settings and explore the timing of adjunctive BI interventions in ED treatment to enhance long-term recovery.

## Body Image Group as Adjunct to Eating Disorder Treatment: Feasibility, Acceptability, and Early Outcomes

Body image (BI) is a multidimensional construct that encompasses the ways in which one perceives, interacts with, thinks, and feels about one’s body (Prnjak et al., 2022). Accordingly, negative BI can involve being dissatisfied with one’s body, holding distorted perceptions of one’s body (e.g., overestimating body size), and judging or evaluating one’s self-worth primarily based on body shape, weight, or overall appearance (Glashouwer et al., 2019). Negative BI can also present behaviorally through the repeated checking or avoidance of one’s body shape or weight (Shafran et al., 2004). Such cognitive, emotional, and behavioral aspects of negative BI are highly prevalent and of significant concern across different cultures (e.g., Frayon et al., 2020; Nikniaz et al., 2016; Rodgers et al., 2023; Santos Silva et al., 2011; Xu et al., 2010), genders (e.g., Baker et al., 2019; Richburg & Stewart, 2024), and other diverse elements of social identity (Frederick et al., 2022; Gordon et al., 2023; Larson et al., 2021). Moreover, youth and adults with negative BI experience impaired physical, sexual, and social health and wellbeing (Haraldstad et al., 2011; Medeiros de Morais et al., 2017; Mond et al., 2013) and are at increased risk for psychopathology and mental health concerns, including depressive symptoms, anxiety disorders, and negative affect (Glashouwer et al., 2019; Sharpe et al., 2018; Vannucci & Ohannessian, 2018; Walker et al., 2018).

Perhaps most notably, negative BI is known to contribute to the development and maintenance of eating disorders (EDs; Gardner et al., 2000; Prnjak et al., 2021; Stice & Shaw, 2002) and is a core component of ED diagnostic criteria and psychopathology (American Psychiatric Association, 2013; Fairburn, 2008). In both clinical and community samples, negative BI positively correlates with maladaptive eating, exercise, and weight-control behaviors (e.g., Brechan & Kvalem, 2015; Grilo et al., 2019; Mitchison et al., 2017; Stutts & Blomquist, 2018). Among individuals with EDs, negative BI is associated with worse treatment outcomes and higher risk of relapse (Boehm et al., 2016; Berends et al., 2018; Pellizzer et al., 2018). Despite these numerous harmful effects, existing ED treatments are minimally effective at ameliorating BI issues. A meta-analysis of randomized controlled trials for anorexia nervosa (AN) found that specialized ED treatments were no more effective at alleviating the psychological symptoms of AN (i.e., body shape and weight concerns) than comparator treatments at both end of treatment and follow up (Murray et al., 2019). Likewise, a transdiagnostic review of the recommended treatment for adults with EDs (enhanced cognitive behavior therapy [CBT-E]; Fairburn, 2008) concluded that although CBT-E holds promise for reducing the overvaluation of body shape and weight in the short term, long-term outcomes are more modest and comparable to comparison treatments (Atwood & Friedman, 2020). Even among youth with EDs (for whom ED prognosis is more favorable than adults; Koreshe et al., 2023), many continue to experience negative BI throughout and following treatment (Le Grange et al., 2019) and thus remain susceptible to the deleterious impact of these symptoms.

Several BI interventions have been developed that directly target negative BI in both ED and community populations using a variety of educational and therapeutic strategies. Alleva and colleagues (2015) conducted a meta-analytic review of stand-alone BI interventions and identified the specific content and techniques that appear to be most effective at improving individuals’ BI. These included CBT-based interventions, as well as those that incorporate discussion of the causes and consequences of negative BI through topics such as media literacy and personal values. Three existing BI interventions across prevention efforts and ED treatment that employ these strategies and have garnered a strong evidence base include the Body Image Workbook (Cash, 2008), the BI module from CBT-E for adolescents with EDs (Dalle Grave & Calugi, 2020), and the Body Project (Stice et al., 2013) and its adaptations (e.g., Ciao et al., 2018). The Body Image Workbook combines psychoeducation, self-monitoring, cognitive restructuring, and relaxation training to address key maintaining factors as conceptualized within the cognitive behavioral model of body image (Cash, 2012), and using the workbook has been found to be effective in reducing negative BI severity among college students (Cash & Hrabosky, 2003; Strachan & Cash, 2002). Extending from this work, CBT-E for adolescents uses primarily behavioral interventions adapted from the adult manual (Fairburn, 2008) to target overvaluation of body shape and weight and the behaviors through which this is expressed (e.g., body checking, body avoidance, marginalization of other areas of life). Multiple clinical trials have found that CBT-E significantly reduces body shape and weight concerns among adults with transdiagnostic EDs (Atwood & Friedman, 2020), and similar findings have been reported for adolescents (Dalle Grave et al., 2021). Finally, as a dissonance-based ED prevention program that focuses on reducing body dissatisfaction by targeting thin-ideal internalization, the Body Project has repeatedly been found to improve BI when delivered to adolescents with BI concerns (see Becker & Stice, 2017 for a review). In one adaptation, the EVERYbody Project (Ciao et al., 2018) incorporated modifications to promote inclusivity among more diverse populations and expand the program’s discussions to include issues of human diversity. The EVERYbody Project supported comparable BI improvements among participants with marginalized and majority identities (Ciao et al., 2021).

Although these evidence-based BI interventions exist, there are minimal guidelines for psychotherapists regarding how to effectively and systematically integrate these resources into existing ED treatments or other psychotherapies to comprehensively address patients’ BI concerns. Clinicians regularly integrate different therapeutic modalities to individualize treatment (Boswell et al., 2019); yet, studies evaluating the effectiveness of these approaches remain limited, and no study to our knowledge has examined the feasibility or effectiveness of adjunctive body image interventions in psychotherapy. Moreover, in order to reach the large proportion of those affected by EDs and negative BI, many of whom have difficulty accessing and benefiting from evidence-based care (Kazdin et al., 2017), any adjunctive BI intervention protocol must be scalable (Cooper & Bailey-Straebler, 2015) and inclusive of all genders and body sizes (Halbeisen et al., 2022; McEntee et al., 2023). Recognizing (1) psychotherapists’ need for more support in addressing negative BI in their patients and (2) systemic barriers to ED services, the authors [KMH and ECA] sought to programmatically incorporate existing BI interventions into an outpatient ED clinic serving adolescents and young adults, with a focus on inclusivity and accessibility. Thus, the aforementioned evidence-based BI interventions were modified to suit a group format that could be delivered virtually, so as to increase access, particularly in the aftermath of the COVID-19 pandemic. As many of these interventions include concepts and exercises that primarily target thin, cisgender females, additional adaptations were made to promote gender inclusivity, weight neutrality, and discussion of the intersectional facets of identity and sociocultural factors that relate to BI (Cafri et al., 2005; Pearl & Puhl, 2016; Tiggemann, 2015).

The current study sought to assess the feasibility and preliminary outcomes of this virtual BI group for adolescents and young adults with transdiagnostic EDs already receiving specialized ED treatment. Specifically, this study evaluated (a) group recruitment, enrollment, and retention; (b) perceived suitability and success of the group by participants both before and after group completion; and (c) improvements in BI and ED pathology over the course of group participation. It was hypothesized that participants would (a) be successfully recruited and retained to participate in and complete an adjunctive BI group intervention; (b) describe the intervention as suitable and successful; and (c) demonstrate reductions in negative BI and ED psychopathology before and after the adjunctive intervention.

## Method

### Participants and Procedure

The BI group protocol was implemented in an outpatient ED clinic at an academic medical center in the United States, where participants were recruited from the Department of Psychiatry and the Adolescent Medicine Division within the Department of Pediatrics. Clinicians within these services referred patients based on the following inclusion criteria: experiencing significant BI concerns and willing and able to participate in group discussions of BI. The initial target age range was 14-18 years old; however, early on in recruitment, this was expanded to include young adults up to age 21 if deemed age-appropriate for the group based on clinical judgment. Patients with current EDs were required to be engaged in outpatient ED treatment (e.g., family-based treatment [FBT], CBT-E) and medically stable. Following guidelines from current evidence-based protocols regarding the timing of BI interventions in ED treatment (Fairburn, 2008; Griffen et al., 2018; Rienecke & Le Grange, 2022), ED patients were also required to be well-nourished (i.e., >90% of expected body weight, in later phases of FBT, based on clinical judgment). To further support their safety, patients were excluded from group participation if they had experienced active suicidal ideation (i.e., with plan or intent), non-suicidal self-injury, or other acute safety concerns in the past two months.

Referred patients first participated in a 30-minute individual session with the primary group facilitator [KMH], who assessed for inclusion/exclusion criteria, provided a brief orientation to the group, and engaged in motivational interviewing techniques (Arkowitz et al., 2015) to facilitate group participation. Data were collected for clinical and quality improvement purposes, and all participants were asked to complete an online survey before starting and, if relevant during the data collection period, after completing or withdrawing from the group. This data on the BI group intervention was then de-identified and retrospectively reviewed for research purposes, with the defined study period spanning the first 18 months of its implementation (March 2022 – August 2023). This study was approved by the authors’ institutional review board with a waiver of informed consent.

### Intervention

The BI group intervention consisted of 12 sessions delivered in three modules. Modules included four sessions and were each based on an evidence-based BI program or intervention, including 1) *Building Acceptance* (based on The Body Project [Stice et al., 2013] and the EVERYBody Project [Ciao et al., 2016]); 2) *Changing Behaviors* (based on CBT-E for adolescents with EDs [Dalle Grave & Calugi, 2020]); and 3) *Cultivating Mindfulness* (based on The Body Image Workbook [Cash, 2008]). Group recruitment was continuous such that new group participants could join at the start of each module, but the group was closed to new participants during the module’s four weeks. Participants were considered to have completed the group when they had attended all three modules.

Each session began with introductions and an ice-breaker question, followed by homework review, teaching and discussion of a new topic, and assigning homework to be completed in the following week. To support skill building and in recognition of the distress that can be elicited by discussing BI-related topics, sessions concluded with a guided mindfulness or grounding exercise drawn from the Dialectical Behavior Therapy (DBT) Skills Manual for Adolescents (Rathus & Miller, 2014) or Seeking Safety (Najavits, 2001).

All group sessions were 60 minutes in length and facilitated via telehealth by one primary facilitator (KMH) and a co-facilitator when available (e.g., another psychologist or psychology learner). Following each group session, group members received an email reminder about the homework and recommendations for additional resources (e.g., books, podcasts, social media accounts) based on that week’s discussion.

## Measures

### Feasibility and Acceptability

The group’s feasibility and acceptability were assessed based on the number of patients who were referred to, screened, enrolled in, and completed the group. To assess the perceived suitability and success of the group, the following questions were posed via the online survey: “How suitable do you think the group is/was for your situation?” and “How successful do you think the group will be/was for you?” Participants rated both questions on a Likert-type scale (0 = “Not at all”; 5 = “Moderate”; 10 = “Extremely”). Text entry boxes were also provided for participants to provide qualitative feedback.

### Eating Disorder Pathology

**Eating Disorder-15 (ED-15).** The ED-15 (Tatham et al., 2015) is a brief measure of ED psychopathology that has been validated in ED clinical samples 13 years and older (Rodrigues et al., 2019). Focusing on the previous week, it includes 10 items that assess ED attitudes, as well as five questions regarding the frequency of ED behaviors. Symptom severity is measured using a 7-point scale (0 = “Not at all”; 6 = “All the time”); the attitudinal items can be averaged together to generate two subscales (Eating Concerns and Weight/Shape Concerns) and a total score. In the current study, Cronbach’s alpha for the total score ranged from .91 to .93 across timepoints.

### Body Image

Participants’ body image was initially assessed (March to November 2022) using the Body Shape Questionnaire (BSQ; Cooper et al., 1987); however, as a clinical tool the BSQ appeared not to be sensitive to change, even among those participants who were endorsing change in BI concerns on the ED-15. Thus, the decision was made to remove the BSQ from the assessment and add three BI questionnaires to capture multiple dimensions of body image: the Sociocultural Attitudes Toward Appearance Questionnaire-4 (SATAQ-4; Schaefer et al., 2015), the Anti-Fat Attitudes Scale (AFAS; Morrison & O’Connor, 1999), and the Body Parts Satisfaction Scale (BPSS; Berscheid et al., 1973).

**Body Shape Questionnaire (BSQ).** The BSQ (Cooper et al., 1987) was designed to assess concerns about body shape and weight. The original measure consists of 34 questions related to distressing preoccupation with one’s weight and shape, body avoidance behaviors, and the experience of “feeling fat.” In responding to the items, individuals are asked to consider how they have been feeling about their appearance over the past four weeks and record their responses on a 6-point scale (1 = “Never”; 6 = “Always”). Items are summed to produce a total score, with higher scores indicating greater body shape concern. The current study used the 8-item version (BSQ-8C; Evans & Dolan, 1993), which has demonstrated the greatest sensitivity to change (Pook et al., 2008). Cronbach’s alpha for the BSQ-8C was .85 at both timepoints.

**Sociocultural Attitudes Toward Appearance Questionnaire-4 (SATAQ-4).** The SATAQ (Heinberg et al., 1995) is a measure of the sociocultural factors that contribute to the acceptance and internalization of appearance ideals. The fourth version of the measure (Schaefer et al., 2015) seeks to provide a comprehensive assessment of potential appearance-related pressures from family, peers, and the media, as well as both thin-ideal and muscular-ideal internalization. Questions are rated on a 5-point scale (1 = “Definitely Disagree”; 5 = “Definitely Agree”); specific items can be averaged together to yield five subscale scores. In the current study, only the 10 questions pertaining to the thin-ideal internalization and muscular/athletic-ideal internalization subscales were used; Cronbach’s alpha for these items ranged from .89 to .90.

**Anti-Fat Attitudes Scale (AFAS).** The AFAS (Morrison & O’Connor, 1999) consists of five items developed to measure negative attitudes toward “fat people.” Respondents indicate the extent to which they agree (1 = “Strongly disagree”; 5 = “Strongly agree”) with five statements about people in larger bodies, with higher scores indicative of stronger anti-fat attitudes.

Cronbach’s alpha in the current study ranged from .65 to .76.

**Body Parts Satisfaction Scale (BPSS).** Nine items from the BPSS (Berscheid et al., 1973) were selected to evaluate participants’ level of satisfaction with nine different body parts or attributes: weight, figure, body build, stomach, waist, thighs, buttocks, hips, and legs. For each body part, satisfaction or dissatisfaction was reported on a 6-point Likert scale (1 = “Extremely satisfied”; 6 = “Extremely dissatisfied”), such that higher scores denoted greater body dissatisfaction. Cronbach’s alpha for the BPSS ranged from .76 to .84.

### Statistical Analyses

All analyses were conducted in SPSS, version 29. Feasibility and acceptability outcomes were analyzed descriptively, including rates of group referral, screening, enrollment, and completion, and the means and standard deviations of questions evaluating group suitability and success. Paired samples *t* tests were used to compare pre- and post-group scores on measures of ED pathology and body image. Cohen’s *d* was reported as a measure of effect size. Only participants who completed assessments at both timepoints were included in pre-post analyses.

## Results

### Feasibility and Acceptability

Of the 61 youth who were referred to the group during the study period, 73.7% (*n* = 45) were screened and 59.0% (*n* = 36) enrolled. The majority of those who declined to enroll in the group after attending a screening visit (*n* = 9) were not interested in the group (77.8%, *n* = 7), while the remainder (22.2%, *n* = 2) were determined to be ineligible at the time of the visit based on their outpatient ED treatment status. Specifically, one ineligible adolescent was less than 90% of their expected body weight and declined medical follow-up, while the other expressed not being “ready” to discuss BI in a group setting.

Demographic information for the final sample (*n* = 36, age range: 14-21) is shown in Table 1. Most participants had AN or atypical AN (69.4%, *n* = 25) and were receiving CBT-E (44.4%, *n* = 16) or FBT (25.0%, *n* = 9) as their primary treatment. Most participants (63.9%, *n* = 23) had completed the group (*n* = 18) or were still active participants in the group (*n* = 5) at the end of the data collection period. About one-third withdrew after starting the group (30.6%, *n* = 11) due to scheduling issues (*n* = 3), needing a higher level of care (*n* = 2) or other outpatient treatment (*n* = 1), feeling too anxious during group (*n* = 2), or perceiving the group as being a poor fit (*n* = 3) due to limited body size and gender diversity across participating members or a personal lack of interest in program content. The remainder were scheduled to participate (5.6%, *n* = 2) at a future entry point.

**Table 1.** Sociodemographic and Clinical Characteristics of Participants.

|  | *M* | *SD* |
| --- | --- | --- |
| Age | 16.19 | 1.47 |
|  | *n* | % |
| Sex |  |  |
| Female | 35 | 97.2 |
| Male | 1 | 2.8 |
| Gender identity |  |  |
| Cisgender female | 31 | 86.1 |
| Non-binary or genderqueer | 3 | 8.3 |
| Cisgender male | 1 | 2.8 |
| Transgender male | 1 | 2.8 |
| Race and ethnicity<sup>a</sup> |  |  |
| White | 18 | 50.0 |
| Asian | 7 | 19.4 |
| Black or African American | 1 | 2.8 |
| Hispanic or Latino | 2 | 5.6 |
| More than one | 7 | 19.4 |
| Unknown | 1 | 2.8 |
| ED diagnosis |  |  |
| Anorexia nervosa | 19 | 52.8 |
| Atypical anorexia nervosa | 6 | 16.7 |
| Other specified feeding or eating disorder | 7 | 19.4 |
| Binge eating disorder | 4 | 11.1 |
| Current primary treatment |  |  |
| CBT-E | 16 | 44.4 |
| FBT | 9 | 25.0 |
| Private practice ED therapy | 6 | 16.7 |
| Other CBT or DBT | 3 | 8.3 |
| Brief Psychology Consultation Clinic<sup>b</sup> | 2 | 5.6 |
| Comorbid mental health diagnosis<sup>c</sup> |  |  |
| One or more | 18 | 50.0 |
| None | 11 | 30.6 |
| Unknown | 7 | 19.4 |
| Frequency of comorbidities<sup>c</sup> |  |  |
| Depression (MDD) | 12 | 33.3 |
| Anxiety (GAD, SAD) | 11 | 30.6 |
| ADHD | 3 | 8.3 |
| OCD | 3 | 8.3 |
| PTSD | 2 | 5.6 |

Note. $N = 36$ . ED = eating disorder; CBT-E = enhanced cognitive behavior therapy; FBT = family-based treatment; CBT = cognitive behavior therapy; DBT = dialectical behavior therapy; MDD = major depressive disorder; GAD = generalized anxiety disorder; ADHD = attention deficit hyperactivity disorder; OCD = obsessive-compulsive disorder; PTSD = post-traumatic stress disorder.
<sup>a</sup> Race and ethnicity were not systematically assessed but reported based on data available in participants' electronic medical records.
<sup>b</sup> The Brief Psychology Consultation Clinic offers three 30-minute psychology sessions for patients awaiting ED-specific behavioral intervention (Bruett et al., 2022).
<sup>c</sup> Comorbid mental health diagnoses reported based on data available in participants' electronic medical records.

Participants who completed the pre-group questionnaire (83.3%, *n* = 30) anticipated that the group would be suitable (*M* = 7.43, *SD* = 1.79) and moderately successful (*M* = 6.17, *SD* = 2.02). At the end of group, 72.2% (*n* = 13) of group completers (*n* = 18) provided post-group data, reporting that the group was suitable (*M* = 7.92, *SD* = 2.50) and successful (*M* = 7.14, *SD* = 2.03) for their needs. In their qualitative feedback, participants described the sense of community and support facilitated by the group. They also indicated that they learned through group discussion and the sharing of one another’s experiences; they also developed helpful coping skills. Constructive feedback included disliking certain group activities or feeling that the group was “too clinical.” Others noted a sense that it would likely take more time than the allotted 12 weeks to experience significant improvements in their BI.

### Eating Disorder Pathology

Descriptive statistics for the ED-15 are shown in Table 2. Among participants who completed the ED-15 both pre- and post-group (*n* = 12), there were significant, large decreases in the Eating Concerns subscale, *t*(10) = 3.81, *p* = .003, *d* = 1.15, Weight and Shape Concerns subscale, *t*(11) = 3.88, *p* = .003, *d* = 1.12, and the total score, *t*(11) = 4.87, *p* < .001, *d* = 1.41, from baseline to end of treatment.

**Table 2.** Descriptive Statistics for Measures of Eating Disorder Pathology and Body Image Before and After Group Participation.

| Measure | Pre-group |  |  | Post-group |  |  |
| --- | --- | --- | --- | --- | --- | --- |
|  | *n* | *M (SD)* | Range | *n* | *M (SD)* | Range |
| ED-15 |  |  |  |  |  |  |
| Eating concerns | 30 | 3.40 (1.42) | 0.75-6.00 | 13 | 2.83 (1.57) | 0.50-5.75 |
| Weight and shape concerns | 30 | 3.97 (1.44) | 0.83-6.00 | 13 | 3.12 (1.32) | 0.50-4.50 |
| Total score | 30 | 3.74 (1.32) | 1.10-6.00 | 13 | 3.02 (1.34) | 0.50-5.00 |
| BSQ | 36 | 16.00 (17.99) | 0.00-48.00 | 36 | 16.00 (17.99) | 0.00-48.00 |
| SATAQ-4 |  |  |  |  |  |  |
| Muscular/athletic internalization | 13 | 3.32 (1.04) | 2.00-5.00 | 6 | 2.53 (1.19) | 1.20-4.60 |
| Thin/low body fat internalization | 13 | 3.99 (0.71) | 2.80-5.00 | 6 | 3.03 (0.78) | 2.00-4.00 |
| Global score | 13 | 3.65 (0.76) | 2.80-4.80 | 6 | 2.72 (0.86) | 1.56-4.22 |
| AFAS | 13 | 1.94 (0.70) | 1.00-3.00 | 6 | 1.87 (0.41) | 1.40-2.60 |
| BPSS | 13 | 3.86 (0.57) | 2.89-5.00 | 6 | 3.46 (0.48) | 2.89-4.22 |

Note. ED-15 = Eating Disorder-15; BSQ = Body Shape Questionnaire; SATAQ-4 = Sociocultural Attitudes Toward Appearance Questionnaire-4; AFAS = Anti-Fat Attitudes Scale; BPSS = Body Parts Satisfaction Scale

### Body Image

Table 2 also reports descriptive statistics for all measures of BI. BSQ scores were unchanged from pre-group to post-group. SATAQ-4, AFAS, and BPSS scores decreased from before to after group participation, although the small number of participants who completed BI measures at both timepoints precluded *t* test analyses.

## Discussion

BI concerns are highly prevalent among adolescents and young adults (McLean et al., 2022; Wang et al., 2019), increasing their risk for ED pathology and impeding ED recovery. The current study added to the limited psychotherapy integration literature by exploring the feasibility, acceptability, and preliminary outcomes of an adjunctive BI group for adolescents receiving other evidence-based treatments. By integrating existing BI interventions into a cohesive protocol delivered in a virtual group format, the BI group met a clinical need for accessible BI support, and was broadly feasible and acceptable to participants. Most referrals successfully enrolled and attended and/or completed the group, suggesting that virtual groups with adolescents are possible and may represent an option for reducing barriers to intervention access and engagement. Moreover, initial findings indicate promise for the effectiveness of this intervention in reducing negative BI and overall ED pathology, which may increase the likelihood of long-lasting ED recovery (Calugi et al., 2018; Glashouwer et al., 2019; Junne et al., 2019; Lock et al., 2013) and benefit overall quality of life (Griffiths et al., 2017). Participants reported feeling supported by the group space and noticing improvements in their knowledge of the different facets of negative BI, the sociocultural factors that influence these domains, and skills that could be used to address these issues. Given the proliferation and refractory nature of negative BI, particularly among youth with EDs, these findings are encouraging and support prior research on the effectiveness of the group format and the incorporated BI interventions.

Consistent with studies of similar group protocols (Ali et al., 2022; Butryn et al., 2014; Ghaderi et al., 2020; Nye & Cash, 2006), almost one third of participants withdrew prior to group completion, pointing to important challenges that can guide future research and intervention design. In addition to scheduling and logistical challenges, adolescents reported not feeling “ready” to discuss BI-related topics due to associated anxiety, ED pathology, or prioritizing other aspects of ED treatment. While some participants expressed feeling more comfortable engaging with the group virtually due to having control over their camera view and less opportunity to engage in body comparisons, others noted that the use of video increased their anxiety surrounding their appearance during group. Additionally, some gender-diverse youth and youth in larger bodies desired greater gender and body size diversity in group composition, which reflects the perspectives of gender-diverse and higher weight individuals in other ED treatment settings (Harrop, 2019; Harrop et al., 2023). Regarding body size in particular, those in larger bodies endorsed feeling that they did not “fit in” with other participants and invalidated by the BI concerns shared by participants in smaller bodies. Other participants noted that some interventions seemed to perpetuate weight stigma; for example, in CBT-E, the assumption that “feeling fat” is always a perceptual distortion and incongruent with one’s actual body size. Prior research has called for modifications to evidence-based ED treatments that account for body size diversity in EDs and the influence of weight stigma (McEntee et al., 2023; Millner & Mulheim, 2021). Such updates may be especially relevant to BI interventions such as this group, which will require more research on the negative BI experiences of individuals in larger bodies with EDs and greater exploration and innovation of concepts and skills that address the needs and risk factors of this population.

Although participants rated the group as successful and reported a significant decrease in ED pathology, ED-15 scores remained elevated, with average total and subscale scores falling above recommended clinical cutoffs (Rodrigues et al., 2019). This finding suggests that body image interventions alone may be helpful but not sufficient for achieving full remission of symptoms; it is also possible that more time is needed to observe the full effects of BI interventions. Moreover, participants’ body shape concerns (as measured by the BSQ) were unchanged from pre-group to post-group. While the 8-item version of the BSQ has previously been found to be sensitive to change over the course of treatment (Pook et al., 2008), its validity and sensitivity have primarily been assessed among adults with bulimia nervosa, who may conceptualize negative BI differently than adolescents with AN-spectrum EDs. As further evidence for this possibility, BSQ scores in the current study were lower than what has been found in prior clinical samples (Warren et al., 2008), potentially highlighting the importance of selecting measures that have been validated more recently as ED diagnostic criteria and clinical populations have evolved.

Descriptively, participants reported fewer anti-fat attitudes and less body ideal internalization (as measured by the SATAQ-4 and AFAS) after completing the group, as well as greater satisfaction with specific parts of their body listed on the BPSS. These improvements are consistent with the content and aspects of BI targeted by the group’s interventions, which focus on education and changing the cognitions and behaviors that influence BI (Lewis-Smith et al., 2019). Additionally, several participants’ primary therapists observed that participants were using skills learned in group to counter diet culture and body talk at home and school, and that they had become more engaged in ED treatment since starting the BI group. Follow-up data in a larger sample are needed to determine if and how the materials and skills shared in the group continue to shift participants’ perspectives and symptoms over time, or affect their engagement in concurrent ED treatments.

This study had several notable limitations that temper its findings and can guide future research in this area. First, as a naturalistic treatment study, its non-experimental design and lack of a control group limits the extent to which improvements in ED pathology and negative BI can be attributed to the BI group itself. Additionally, because all participants were concurrently involved in individual ED treatment, it is possible that the primary treatment, rather than the BI group, or interactions between the two interventions, caused changes in participants’ symptoms. These findings must also be considered within the context of the small sample size and poor rate of questionnaire completion, particularly post-treatment, and especially for the BI questionnaires. Future research can build on this study’s findings using randomized designs or experimental methodologies to study intervention effects on BI with larger participant samples.

This study was also constrained to a specialized ED clinic within an academic medical center and specific referral criteria; thus, additional research is needed to determine the feasibility and acceptability of this group in other treatment settings and among other treatment-seeking samples. Given the mixed feedback from participants regarding the virtual group format, future research can include consideration of whether some adolescents might be better engaged through an in-person group. Additionally, as participation in the current BI group was limited to adolescents in the later stages of ED treatment (e.g., >90% of expected body weight, in later phases of FBT), future studies can explore the timing of adjunctive BI interventions and at what point in the course of ED recovery they are most likely to be beneficial. Current ED treatment protocols and BI interventions recommend addressing low weight before BI concerns so as not to distract from necessary efforts to increase nutrition and weight (Rienecke & Le Grange, 2022) or inadvertently encourage patients to habituate to body weights/shapes that are medically dangerous (Griffen et al., 2018). Moreover, malnutrition may maintain or exacerbate negative BI in adolescents with EDs (Griffiths et al., 2021) and therefore may be an important first target in the treatment of BI concerns. However, particularly given the limited effectiveness of existing ED treatments in reducing negative BI (Atwood & Friedman, 2020; Le Grange et al., 2019; Murray et al., 2019), future studies might explore whether the implementation of BI interventions earlier in the course of treatment might lead to better BI and weight outcomes.

Finally, most referrals and participants were White, cisgender females with AN. Of note, due to longstanding, inaccurate ED stereotypes and structural barriers to ED services, this demographic is often the primary group presenting to many ED treatments, highlighting a significant issue in the ED field (Halbeisen et al., 2022). As experiences of discrimination and other structural and systemic forms of oppression may differentially influence ED symptoms, case identification, and treatment (Burke et al., 2020), increased efforts are urgently needed to better capture the experiences of minoritized youth receiving ED services (including adjunctive and BI interventions) to improve treatment access and effectiveness for all adolescents with EDs.

Negative BI may be one of the most difficult aspects of ED pathology to address (Atwood & Friedman, 2020; Le Grange et al., 2019; Murray et al., 2019) and often requires long-term and focused efforts on the part of patients and psychotherapists to see change. During adolescence, a critical period of BI development (Wang et al., 2019), negative BI can take many forms and impact functioning and pathology to varying degrees. The BI group examined in the current study provides a model for integrating a range of existing BI interventions, mindfulness, and distress tolerance exercises, thereby offering many opportunities for participants’ unique symptoms and experiences to be discussed. Although preliminary findings support the implementation of this group to bolster the effectiveness of individual ED treatment, additional research is necessary to evaluate the group’s feasibility in other treatment settings, comprehensively assess its effectiveness, and improve the reach and inclusiveness of its recruitment and content.

## Ethics Approval and Consent to Participate

This study was approved by the University of California, San Francisco’s Institutional Review Board (IRB) with a waiver of informed consent and exemption from continuing review because it did not meet the definition of human subjects research.

## Data Availability

All data produced in the present study are available upon reasonable request to the authors.

## References

Ali, K., Fassnacht, D. B., Farrer, L. M., Rieger, E., Moessner, M., Bauer, S., & Griffiths, K. M. (2022). Recruitment, adherence and attrition challenges in internet-based indicated prevention programs for eating disorders: Lessons learned from a randomised controlled trial of ProYouth OZ. Journal of Eating Disorders, 10(1), 1. 10.1186/s40337-021-00520-7

Alleva, J. M., Sheeran, P., Webb, T. L., Martijn, C., & Miles, E. (2015). A meta-analytic review of stand-alone interventions to improve body image. Plos One, 10(9), e0139177-. 10.1371/journal.pone.0139177

American Psychiatric Association. (2013). Diagnostic and statistical manual of mental disorders (5th ed.). 10.1176/appi.books.9780890425596

Arkowitz, H., Miller, W. R., & Rollnick, S. (Eds.). (2015). Motivational interviewing in the treatment of psychological problems (2nd ed.). Guilford Press.

Atwood, M. E., & Friedman, A. (2020). A systematic review of enhanced cognitive behavioral therapy (CBT-E) for eating disorders. International Journal of Eating Disorders, 53(3), 311–330. 10.1002/eat.23206

Baker, J. H., Higgins Neyland, M. K., Thornton, L. M., Runfola, C. D., Larsson, H., Lichtenstein, P., & Bulik, C. (2019). Body dissatisfaction in adolescent boys. Developmental Psychology, 55(7), 1566–1578. 10.1037/dev0000724

Becker, C. B., & Stice, E. (2017). From efficacy to effectiveness to broad implementation: Evolution of the body project. Journal of Consulting and Clinical Psychology, 85(8), 767–782. 10.1037/ccp0000204

Berends, T., Boonstra, N., & van Elburg, A. (2018). Relapse in anorexia nervosa: A systematic review and meta-analysis. Current Opinion in Psychiatry, 31(6), 445–455. 10.1097/YCO.0000000000000453

Berscheid, E., Walster, E., & Bohrnstedt, G. (1973). The happy American body: A survey report. Psychology Today, 7, 119–131.

Boehm, I., Finke, B., Tam, F. I., Fittig, E., Scholz, M., Gantchev, K., Roessner, V., & Ehrlich, S. (2016). Effects of perceptual body image distortion and early weight gain on long-term outcome of adolescent anorexia nervosa. European Child & Adolescent Psychiatry, 25(12), 1319–1326. 10.1007/s00787-016-0854-1

Boswell, J. F., Newman, M. G., & McGinn, L. K. (2019). Outcome research on psychotherapy integration. In J. C. Norcross & M. R. Goldfried (Eds.), Handbook of psychotherapy integration (3rd ed., pp. 405-431). Oxford University Press. 10.1093/medpsych/9780190690465.003.0019

Brechan, I., & Kvalem, I. L. (2015). Relationship between body dissatisfaction and disordered eating: Mediating role of self-esteem and depression. Eating Behaviors, 17, 49–58. 10.1016/j.eatbeh.2014.12.008

Bruett, L. D., Forsberg, S., Accurso, E. C., Gorrell, S., Hail, L., Keyser, J., Le Grange, D., & Huryk, K. M. (2022). Development of evidence-informed bridge programming to support an increased need for eating disorder services during the COVID-19 pandemic. Journal of Eating Disorders, 10(1), 71. 10.1186/s40337-022-00590-1

Burke, N. L., Schaefer, L. M., Hazzard, V. M., & Rodgers, R. F. (2020). Where identities converge: The importance of intersectionality in eating disorders research. International Journal of Eating Disorders, 53(10), 1605–1609. 10.1002/eat.23371

Butryn, M. L., Rohde, P., Marti, C. N., & Stice, E. (2014). Do participant, facilitator, or group factors moderate effectiveness of the Body Project? Implications for dissemination. Behaviour Research and Therapy, 61, 142–149. 10.1016/j.brat.2014.08.004

Cafri, G., Yamamiya, Y., Brannick, M., & Thompson, J. K. (2005). The influence of sociocultural factors on body image: A meta-analysis. Clinical Psychology: Science and Practice, 12(4), 421–433. 10.1093/clipsy.bpi053

Calugi, S., El Ghoch, M., Conti, M., & Dalle Grave, R. (2018). Preoccupation with shape or weight, fear of weight gain, feeling fat and treatment outcomes in patients with anorexia nervosa: A longitudinal study. Behaviour Research and Therapy, 105, 63–68. 10.1016/j.brat.2018.04.001

Cash, T. F. (2008). The body image workbook: An 8-step program for learning to like your looks (2nd ed.). New Harbinger Publications.

Cash, T. F. (2012). Cognitive-behavioral perspectives on body image. In T. F. Cash (Ed.), Encyclopedia of body image and human appearance (pp. 334–342). Elsevier Academic Press.

Cash, T. F., & Hrabosky, J. I. (2003). The effects of psychoeducation and self-monitoring in a cognitive-behavioral program for body-image improvement. Eating Disorders, 11(4), 255–270. 10.1080/10640260390218657

Ciao, A. C., Munson, B. R., Pringle, K. D., Roberts, S. R., Lalgee, I. A., Lawley, K. A., & Brewster, J. (2021). Inclusive dissonance-based body image interventions for college students: Two randomized-controlled trials of the EVERYbody Project. Journal of Consulting and Clinical Psychology, 89(4), 301–315. 10.1037/ccp0000636

Ciao, A. C., Ohls, O. C., & Pringle, K. D. (2018). Should body image programs be inclusive? A focus group study of college students. International Journal of Eating Disorders, 51(1), 82–86. 10.1002/eat.22794

Cooper, P. J., Taylor, M. J., Cooper, Z., & Fairbum, C. G. (1987). The development and validation of the Body Shape Questionnaire. International Journal of Eating Disorders, 6(4), 485–494. 10.1002/1098-108X(198707)6:4<485::AID-EAT2260060405>3.0.CO;2-O

Cooper, Z., & Bailey-Straebler, S. (2015). Disseminating evidence-based psychological treatments for eating disorders. Current Psychiatry Reports, 17(3), 12. 10.1007/s11920-015-0551-7

Dalle Grave, R., & Calugi, S. (2020). Cognitive behavior therapy for adolescents with eating disorders. Guilford Press.

Dalle Grave, R., Conti, M., Sartirana, M., Sermattei, S., & Calugi, S. (2021). Enhanced cognitive behaviour therapy for adolescents with eating disorders: A systematic review of current status and future perspectives. IJEDO, 3, 1–11. 10.32044/ijedo.2021.01

Evans, C., & Dolan, B. (1993). Body Shape Questionnaire: Derivation of shortened “alternate forms”. International Journal of Eating Disorders, 13(3), 315–321. 10.1002/1098-108x(199304)13:3<315::aid-eat2260130310>3.0.co;2-3

Fairburn, C. G. (2008). Cognitive behavior therapy and eating disorders. Guilford Press.

Frayon, S., Cavaloc, Y., Wattelez, G., Cherrier, S., Touitou, A., Zongo, P., Yacef, K., Caillaud, C., Lerrant, Y., & Galy, O. (2020). Body image, body dissatisfaction and weight status of Pacific adolescents from different ethnic communities: A cross-sectional study in New Caledonia. Ethnicity & Health, 25(2), 289–304. 10.1080/13557858.2017.1398818

Frederick, D. A., Crerand, C. E., Brown, T. A., Perez, M., Best, C. R., Cook-Cottone, C. P., Compte, E. J., Convertino, L., Gordon, A. R., Malcarne, V. L., Nagata, J. M., Parent, M. C., Pennesi, J. L., Pila, E., Rodgers, R. F., Schaefer, L. M., Thompson, J. K., Tylka, T. L., & Murray, S. B. (2022). Demographic predictors of body image satisfaction: The US Body Project I. Body Image, 41, 17–31. 10.1016/j.bodyim.2022.01.011

Gardner, R. M., Stark, K., Friedman, B. N., & Jackson, N. A. (2000). Predictors of eating disorder scores in children ages 6 through 14: A longitudinal study. Journal of Psychosomatic Research, 49(3), 199–205. 10.1016/s0022-3999(00)00172-0

Ghaderi, A., Stice, E., Andersson, G., Enö Persson, J., & Allzén, E. (2020). A randomized controlled trial of the effectiveness of virtually delivered Body Project (vBP) groups to prevent eating disorders. Journal of Consulting and Clinical Psychology, 88(7), 643–656. 10.1037/ccp0000506

Glashouwer, K. A., van der Veer, R. M. L., Adipatria, F., de Jong, P. J., & Vocks, S. (2019). The role of body image disturbance in the onset, maintenance, and relapse of anorexia nervosa: A systematic review. Clinical Psychology Review, 74, 101771. 10.1016/j.cpr.2019.101771

Gordon, A. R., Beccia, A. L., Egan, N., & Lipson, S. K. (2024). Intersecting gender identity and racial/ethnic inequities in eating disorder risk factors, symptoms, and diagnosis among US college students: An intersectional multilevel analysis of individual heterogeneity and discriminatory accuracy. International Journal of Eating Disorders, 57(1), 146–161. 10.1002/eat.24089

Griffen, T. C., Naumann, E., & Hildebrandt, T. (2018). Mirror exposure therapy for body image disturbances and eating disorders: A review. Clinical Psychology Review, 65, 163–174. 10.1016/j.cpr.2018.08.006

Griffiths, K. R., Martin Monzon, B., Madden, S., Kohn, M. R., Touyz, S., Sachdev, P. S., Clarke, S., Foroughi, N., & Hay, P. (2021). White matter microstructural differences in underweight adolescents with anorexia nervosa and a preliminary longitudinal investigation of change following short-term weight restoration. Eating and Weight Disorders, 26(6), 1903–1914. 10.1007/s40519-020-01041-z

Griffiths, S., Murray, S. B., Bentley, C., Gratwick-Sarll, K., Harrison, C., & Mond, J. M. (2017). Sex differences in quality of life impairment associated with body dissatisfaction in adolescents. Journal of Adolescent Health, 61(1), 77–82. 10.1016/j.jadohealth.2017.01.016

Grilo, C. M., Crosby, R. D., & Machado, P. P. P. (2019). Examining the distinctiveness of body image concerns in patients with anorexia nervosa and bulimia nervosa. International Journal of Eating Disorders, 52(11), 1229–1236. 10.1002/eat.23161

Halbeisen, G., Brandt, G., & Paslakis, G. (2022). A plea for diversity in eating disorders research. Frontiers in Psychiatry, 13. 10.3389/fpsyt.2022.820043

Haraldstad, K., Christophersen, K. A., Eide, H., Nativg, G. K., & Helseth, S. (2011). Predictors of health-related quality of life in a sample of children and adolescents: A school survey. Journal of Clinical Nursing, 20(21-22), 3048–3056. 10.1111/j.1365-2702.2010.03693.x

Harrop E. N. (2019). Typical-atypical interactions: One patient’s experience of weight bias in an inpatient eating disorder treatment setting. Women & Therapy, 42(1-2), 45–58. 10.1080/02703149.2018.1524068

Harrop, E. N., Hecht, H. K., Harner, V., Call, J., & Holloway, B. T. (2023). “How do I exist in this body…that’s outside of the norm?” Trans and nonbinary experiences of conformity, coping, and connection in atypical anorexia. International Journal of Environmental Research and Public Health, 20(2), 1156. 10.3390/ijerph20021156

Heinberg, L. J., Thompson, J. K., & Stormer, S. (1995). Development and validation of the Sociocultural Attitudes Towards Appearance Questionnaire. International Journal of Eating Disorders, 17(1), 81–89. 10.1002/1098-108x(199501)17:1<81::aid-eat2260170111>3.0.co;2-y

Junne, F., Wild, B., Resmark, G., Giel, K. E., Teufel, M., Martus, P., Ziser, K., Friederich, H. C., de Zwaan, M., Löwe, B., Dinkel, A., Herpertz, S., Burgmer, M., Tagay, S., Rothermund, E., Zeeck, A., Herzog, W., & Zipfel, S. (2019). The importance of body image disturbances for the outcome of outpatient psychotherapy in patients with anorexia nervosa: Results of the ANTOP-study. European Eating Disorders Review, 27(1), 49–58. 10.1002/erv.2623

Kazdin, A. E., Fitzsimmons-Craft, E. E., & Wilfley, D. E. (2017). Addressing critical gaps in the treatment of eating disorders. International Journal of Eating Disorders, 50(3), 170–189. 10.1002/eat.22670

Koreshe, E., Paxton, S., Miskovic-Wheatley, J., Bryant, E., Le, A., Maloney, D., National Eating Disorder Research Consortium, Touyz, S., & Maguire, S. (2023). Prevention and early intervention in eating disorders: Findings from a rapid review. Journal of Eating Disorders, 11(1), 38. 10.1186/s40337-023-00758-3

Larson, N., Loth, K., Eisenberg, M., Hazzard, V., & Neumark-Sztainer, D. (2021). Body dissatisfaction and disordered eating are prevalent problems among US young people from diverse socioeconomic backgrounds: Findings from the EAT 2010–2018 study. Eating Behaviors, 42, 101535. 10.1016/j.eatbeh.2021.101535.

Le Grange, D., Huryk, K. M., Murray, S. B., Hughes, E. K., Sawyer, S. M., & Loeb, K. L. (2019). Variability in remission in family therapy for anorexia nervosa. International Journal of Eating Disorders, 52(9), 996–1003. 10.1002/eat.23138

Lewis-Smith, H., Diedrichs, P. C., & Halliwell, E. (2019). Cognitive-behavioral roots of body image therapy and prevention. Body Image, 31, 309–320. 10.1016/j.bodyim.2019.08.009

Lock, J. D., Agras, W. S., Grange, D. L., Couturier, J. L., Safer, D. L., & Bryson, S. W. (2013). Do end of treatment assessments predict outcome at follow-up in eating disorders? International Journal of Eating Disorders, 46 *8*, 771–778. 10.1002/eat.22175

McEntee, M. L., Philip, S. R., & Phelan, S. M. (2023). Dismantling weight stigma in eating disorder treatment: Next steps for the field. Frontiers in Psychiatry, 14. 10.3389/fpsyt.2023.1157594

McLean, S. A., Rodgers, R. F., Slater, A., Jarman, H. K., Gordon, C. S., & Paxton, S. J. (2022). Clinically significant body dissatisfaction: Prevalence and association with depressive symptoms in adolescent boys and girls. European Child & Adolescent Psychiatry, 31(12), 1921–1932. 10.1007/s00787-021-01824-4

Medeiros de Morais, M. S., Andrade do Nascimento, R., Vieira, M. C. A., Moreira, M. A., Câmara, S. M. A. D., Campos Cavalcanti Maciel, Á., & Almeida, M. D. G. (2017). Does body image perception relate to quality of life in middle-aged women?. PloS One, 12(9), e0184031. 10.1371/journal.pone.0184031

Millner, R., & Mulheim, L. (2021). Anti-fat bias in evidence-based psychotherapies for eating disorders. Can they be adapted to address the harm? In H. Brown & N. Ellis-Ordway (Eds.), Weight bias in health education: Critical perspectives for pedagogy and practice. Routledge. 10.4324/9781003057000-14

Mitchison, D., Hay, P., Griffiths, S., Murray, S. B., Bentley, C., Gratwick-Sarll, K., Harrison, C., & Mond, J. (2017). Disentangling body image: The relative associations of overvaluation, dissatisfaction, and preoccupation with psychological distress and eating disorder behaviors in male and female adolescents. International Journal of Eating Disorders, 50(2), 118–126. 10.1002/eat.22592

Mond, J., Mitchison, D., Latner, J., Hay, P., Owen, C., & Rodgers, B. (2013). Quality of life impairment associated with body dissatisfaction in a general population sample of women. BMC Public Health, 13, 920. 10.1186/1471-2458-13-920

Morrison, T. G., & O’Connor, W. E. (1999). Psychometric properties of a scale measuring negative attitudes toward overweight individuals. Journal of Social Psychology, 139(4), 436–445. 10.1080/00224549909598403

Murray, S. B., Quintana, D. S., Loeb, K. L., Griffiths, S., & Le Grange, D. (2019). Treatment outcomes for anorexia nervosa: a systematic review and meta-analysis of randomized controlled trials. Psychological Medicine, 49(4), 535–544. 10.1017/S0033291718002088

Najavits, L. M. (2001). Seeking safety: A treatment manual for PTSD and substance abuse. Guilford Press.

Nikniaz, Z., Mahdavi, R., Amiri, S., Ostadrahimi, A., & Nikniaz, L. (2016). Factors associated with body image dissatisfaction and distortion among Iranian women. Eating Behaviors, 22, 5–9. 10.1016/j.eatbeh.2016.03.018

Nye, S., & Cash, T. F. (2006). Outcomes of manualized cognitive-behavioral body image therapy with eating disordered women treated in a private clinical practice. Eating Disorders, 14(1), 31–40. 10.1080/10640260500403840

Pearl, R. L., & Puhl, R. M. (2016). The distinct effects of internalizing weight bias: An experimental study. Body Image, 17, 38–42. 10.1016/j.bodyim.2016.02.002

Pellizzer, M. L., Waller, G., & Wade, T. D. (2018). Body image flexibility: A predictor and moderator of outcome in transdiagnostic outpatient eating disorder treatment. International Journal of Eating Disorders, 51(4), 368–372. 10.1002/eat.22842

Pook, M., Tuschen-Caffier, B., & Brähler, E. (2008). Evaluation and comparison of different versions of the Body Shape Questionnaire. Psychiatry Research, 158(1), 67–73. 10.1016/j.psychres.2006.08.002

Prnjak, K., Hay, P., Mond, J., Bussey, K., Trompeter, N., Lonergan, A., & Mitchison, D. (2021). The distinct role of body image aspects in predicting eating disorder onset in adolescents after one year. Journal of Abnormal Psychology, 130(3), 236–247. 10.1037/abn0000537

Rathus, J. H., & Miller, A. L. (2014). DBT skills manual for adolescents. Guilford Press.

Richburg, A., & Stewart, A. J. (2024). Body image among sexual and gender minorities: An intersectional analysis. Journal of Homosexuality, 71(2), 319–343. 10.1080/00918369.2022.2114399

Rienecke, R.D., & Le Grange, D. (2022). The five tenets of family-based treatment for adolescent eating disorders. Journal of Eating Disorders, 10, 60. 10.1186/s40337-022-00585-y

Rodgers, R. F., Laveway, K., Campos, P., & de Carvalho, P. H. B. (2023). Body image as a global mental health concern. Global Mental Health, 10, e9. 10.1017/gmh.2023.2

Rodrigues, T., Vaz, A. R., Silva, C., Conceição, E., & Machado, P. P. P. (2019). Eating Disorder-15 (ED-15): Factor structure, psychometric properties, and clinical validation. European Eating Disorders Review, 27(6), 682–691. 10.1002/erv.2694

Santos Silva, D. A., Nahas, M. V., de Sousa, T. F., Del Duca, G. F., & Peres, K. G. (2011). Prevalence and associated factors with body image dissatisfaction among adults in southern Brazil: A population-based study. Body Image, 8(4), 427–431. 10.1016/j.bodyim.2011.05.009

Schaefer, L. M., Burke, N. L., Thompson, J. K., Dedrick, R. F., Heinberg, L. J., Calogero, R. M., Bardone-Cone, A. M., Higgins, M. K., Frederick, D. A., Kelly, M., Anderson, D. A., Schaumberg, K., Nerini, A., Stefanile, C., Dittmar, H., Clark, E., Adams, Z., Macwana, S., Klump, K. L., Vercellone, A. C., … Swami, V. (2015). Development and validation of the Sociocultural Attitudes Towards Appearance Questionnaire-4 (SATAQ-4). Psychological Assessment, 27(1), 54–67. 10.1037/a0037917

Shafran, R., Fairburn, C. G., Robinson, P., & Lask, B. (2004). Body checking and its avoidance in eating disorders. International Journal of Eating Disorders, 35(1), 93–101. 10.1002/eat.10228

Sharpe, H., Patalay, P., Choo, T. H., Wall, M., Mason, S. M., Goldschmidt, A. B., & Neumark-Sztainer, D. (2018). Bidirectional associations between body dissatisfaction and depressive symptoms from adolescence through early adulthood. Development and Psychopathology, 30(4), 1447–1458. 10.1017/S0954579417001663

Stice, E., Rohde, P., & Shaw, H. (2013). The body project: A dissonance-based eating disorder prevention intervention (2nd ed.). Oxford University Press.

Stice, E., & Shaw, H. E. (2002). Role of body dissatisfaction in the onset and maintenance of eating pathology: A synthesis of research findings. Journal of Psychosomatic Research, 53(5), 985–993. 10.1016/s0022-3999(02)00488-9

Strachan, M. D., & Cash, T. F. (2002). Self-help for a negative body image: A comparison of components of a cognitive-behavioral program. Behavior Therapy, 33(2), 235–251. 10.1016/S0005-7894(02)80027-2

Tiggemann M. (2015). Considerations of positive body image across various social identities and special populations. Body Image, 14, 168–176. 10.1016/j.bodyim.2015.03.002

Vannucci, A., & Ohannessian, C. M. (2018). Body image dissatisfaction and anxiety trajectories during adolescence. Journal of Clinical Child and Adolescent Psychology, 47(5), 785– 795. 10.1080/15374416.2017.1390755

Walker, D. C., White, E. K., & Srinivasan, V. J. (2018). A meta-analysis of the relationships between body checking, body image avoidance, body image dissatisfaction, mood, and disordered eating. International Journal of Eating Disorders, 51(8), 745–770. 10.1002/eat.22867

Wang, S. B., Haynos, A. F., Wall, M. M., Chen, C., Eisenberg, M. E., & Neumark-Sztainer, D. (2019). Fifteen-year prevalence, trajectories, and predictors of body dissatisfaction from adolescence to middle adulthood. Clinical Psychological Science, 7(6), 1403–1415. 10.1177/2167702619859331

Warren, C. S., Cepeda-Benito, A., Gleaves, D. H., Moreno, S., Rodriguez, S., Fernandez, M. C., Fingeret, M. C., & Pearson, C. A. (2008). English and Spanish versions of the Body Shape Questionnaire: Measurement equivalence across ethnicity and clinical status. International Journal of Eating Disorders, 41(3), 265–272. 10.1002/eat.20492

Xu, X., Mellor, D., Kiehne, M., Ricciardelli, L. A., McCabe, M. P., & Xu, Y. (2010). Body dissatisfaction, engagement in body change behaviors and sociocultural influences on body image among Chinese adolescents. Body Image, 7(2), 156–164. 10.1016/j.bodyim.2009.11.003

